# “He stepped on my belly” An exploration of Intimate Partner Violence Experience and Coping Strategies among Pregnant Women in Southwestern-Uganda

**DOI:** 10.1101/2023.10.25.23297494

**Authors:** Eve Katushabe, JohnBosco Ndinawe, Abeneitwe Editor, Katusiime Agnes, Gladys Nakidde, John Baptist Asiimwe, Vincent Batwala

## Abstract

Intimate Partner Violence (IPV) associated with pregnancy remains a challenge globally and in the Ugandan context. However, literature on IPV experiences, support seeking and coping strategies during pregnancy remains limited in Uganda. This study explored the pregnant women’s IPV experiences, support seeking and coping strategies in Southwestern Uganda. Pregnant women with IPV experience during the index pregnancy were purposively approached for in-depth interviews and saturation of data was reached at 25 respondents. Data was analyzed inductively using thematic analysis.

Women voiced experiences of IPV that included partners: spending nights away from home without any communication, refusal of accompaniment for antenatal care contacts, uncomfortable sexual intercourse positions, forced sexual intercourse, being slapped, punched, and kicked, failure to pay bills like rent, children’s school fees, transport money to seek medical care and food. Women preferred sharing IPV experiences with their biological mothers to midwives or any other person and some kept it to themselves. The main support given by their support systems was encouraging the victims to try and maintain their marriage and keeping quiet when the partner starts quarreling. Women coped by confiding in their relatives, keeping silent, self-consolation, tolerance of the perpetrator since they financially depended on them, distracting bad thoughts through thinking about good things like friends, self-blame and praying to God. Pregnant women did not understand the role of midwives in IPV nor did the midwives’ inquire about the IPV experience during Antenatal care contacts.

The findings of this study point to the need for the Health system to incorporate a user friendly IPV screening tool onto the ANC card to enhance routine IPV screening by midwives and recruit counselors and peer supporters to assist midwives in providing individualized psychological support.

## Introduction

Intimate partner violence (IPV) is a public health problem and has been recognized as a serious social issue^(1, 2)^ which affects all women irrespective of cultural or religious affiliation, socioeconomic and education background^(3, 4)^. IPV manifests in forms of physical, sexual, psychological^(5, 6)^ and economic abuse ^(7, 8)^. In addition to other forms of violence, economic violence has been identified more recently as another form of IPV. It involves behaviors that control the victim’s power to get, use and maintain resources hence lowering the victim’s economic security and possibility for self-sufficiency^(9)^. Furthermore, economic violence affects both the woman and children since it contributes to home instabilities especially when the perpetrators neglect their role as family breadwinners which is contrary to the African culture(10, 11). Globally, it is known that one in three women experience IPV in their life time, with African countries ranking highest^(12)^ and reporting IPV in pregnancy at 66.9% in Kenya, 18.8% in Tanzania, 41.1%, 59% in Ethiopia and 61.8% in Gambia^(13–17)^. In Uganda among women of the reproductive age (15-49 years), 40% of the ever married experienced IPV ^(18)^ and according to the Uganda Bureau of Statistics (UBOS), perinatal IPV was reported at 10.6%, while IPV during the index pregnancy at 27.8% in Eastern Uganda and 70.3% in Southwestern Uganda ^(18–20)^.

Occurrence of IPV negatively affects pregnancy outcomes for both mother and infant ^(21, 22)^. Earlier studies reported that women experiencing IPV during pregnancy are at a higher risk of chronic stress^(23)^, spontaneous abortion, delayed antenatal care first contact, sexually transmitted diseases acquisition, antepartum hemorrhage, trauma related to IPV, feeling of defenselessness and other health problems ^(18, 24–28)^. These may lead to serious mental health problems like substance and alcohol abuse, depression and suicidal tendencies ^(25, 28–32)^. Effects on the baby include: premature rupture of membranes, risk of preterm birth, low birth weight, fetal injuries, perinatal and neonatal mortality ^(25, 28, 33, 34)^. Pregnant women experience shame, fear and stigma following IPV episodes, which significantly contributes to victim isolation and concealment of her experiences yet involving other people can be a strategy to cope with the violence^(35, 36)^. nevertheless some studies indicate that a number of victims will seek support in severe IPV cases^(37)^, which helps them in dealing with the traumatizing life events ^(38)^. Studies report that IPV victims adapting certain coping strategies have lower risks of suffering from alexithymia and depression^(39, 40)^. The identified coping strategies include: using various distracters to avoid bad feelings like watching movies, singing and talking to friends averting their intimate partners’ attention and protesting against violence^(38, 41)^, and seeking social support, problem solving and self-re-examination, dedication to religion which involves praying, reading the Bible and visiting pastors for counseling and prayers^(42–45)^. Some victims adopt coping strategies like fighting back or shaming the partner by disclosing the violent behavior beyond family members to co-workers or friends, self-blame ^(43, 44)^, acceptance, using emotion-focused strategies which include substance use (including alcohol, other drugs) ^(43)^. Coping strategies commonly in African settings include: self-re-examination, devotion to religion and quest for informal social support. These practices can positively or negatively impact onto ones’ mental or physical health^(41)^.

Concentrating on coping strategies used by women can be helpful in developing focused IPV management interventions^(46)^. Pregnancy provides a unique opportunity for health care providers to connect with women in time enabling identification of IPV experiences and its distinct effects ^(47)^, as they come for ANC contacts. Currently women in Uganda are recommended to have eight ^(8)^ contacts with care providers during pregnancy and IPV screening is an important ANC package for every woman ^(48)^. Though the Uganda clinical guidelines (UCG) recommends IPV screening during the prenatal period^(49)^, there is a paucity of documented information regarding qualitative inquiry of pregnant women’s IPV experiences and their coping strategies.

Many studies have reported IPV experience, prevalence and associated factors during pregnancy using quantitative methods ^(6, 19)^. Therefore, our aim was to explore IPV experiences, coping and support seeking strategies among pregnant women attending a city hospital in Southwestern Uganda.

## Materials and Methods

This study was carried out within a larger study titled “Intimate partner violence disclosure among pregnant women attending a health facility in South Western Uganda” An interpretive phenomenological qualitative study design was employed to explore pregnant women’s IPV experiences, support seeking and their coping strategies.

The study was conducted at the ANC clinic of a high-volume City hospital for a period of one month from 7^th^ January 2019 to 7^th^ February 2019. The hospital operates a daily general outpatient, Antenatal, Family planning and Young Child Clinics; and an inpatient Maternity Ward where normal deliveries, assisted and operative deliveries are conducted. The facility ANC monthly attendance was 800 (including new and re-attendance) as per 2018 Hospital database (both new ANC cases and re-attendance). These ANC consultations for all low risk pregnant women are managed by midwives on a daily basis Monday to Friday excluding Thursday which is strictly for pregnant women living with Human Immune Virus (HIV) and high risk cases are managed by the medical officer with room for referral to the nearby Regional Referral Hospital. Participants aged 15 and above in a larger cohort of 199 pregnant women that had experienced at least one form of IPV were enrolled for the in depth interviews.

Purposeful sampling ^(50)^ was used to select pregnant women that had experienced IPV. The sample size was a maximum of 25 respondents; this was guided by the saturation of data attained during the in-depth interviews.

An In-depth interview guide was used. We pre-tested the interview guide on five pregnant women attending ANC from a different health facility. Ambiguous questions in the guide were amended for clarity before data collection commenced. The interview guide contained questions on the bio-demographic characteristics (Age, tribe, level of education), probing questions about the type of IPV experienced “What form of intimate partner violence (IPV) have you experienced during this current pregnancy?” How did you feel when this was experienced?”:“What did you do when this happened?”:“Why did you do that?”:“Have you ever told any midwife voluntarily about the IPV experience?”: “Have you ever been asked by a midwife about IPV during pregnancy”: “What do you do to cope with the violent partner? “The interview guide was informed by literature on IPV ^(13, 37, 43, 51)^.

We carried out in-depth interviews which were audio recorded and field notes were taken. Each interview took between 45 to 60 minutes. The interviews were majorly carried out in the local language Runyankole to ensure the best quality of data since it is the commonly used language in the area.

Interviews were conducted by KE who is experienced in collecting qualitative data and two baccalaureate nurses who were trained in qualitative data collection.

The first author and interviewer introduced the study to the health facility in-charge and other health care providers in person to introduce the study and create rapport. Participants were informed that participation is voluntary and decision to withdraw wouldn’t affect their routine care. Recruitment was done at the exit after receiving all the ANC services. In-depth interviews were carried out in the ANC clinic private rooms which prevented interruptions and supported privacy. Before each interview session, written informed consent was obtained. Data identifiers were not considered to maintain confidentiality and data safety, therefore pseudonyms or false names have been used in this study.

Data was analyzed using thematic analysis approach. Thematic analysis was selected for the study as it allows analysis and understanding of the meaning of various coded responses from interviews in their right context^(52)^. Initially interviews were transcribed in Runyankore and then translated into English. Transcription and translation were done by KE and NJB and these checked themselves. The translated scripts were repetitively read by three researchers individually to familiarize themselves to the data and pinpoint significant responses connected to the study objectives as well as distinctive responses to the prompts consequently inductive coding was done. After finding repetitive expressions, responses that included the expressions were highlighted and color coded in each interview. The color coded phrases were grouped into categories and the categories structured into themes. During this analysis, themes directly connected to the study objectives were identified. In addition, emergent and new themes were also established. The themes were revised by the team for accuracy and to authenticate that they represented the correct description of the respondents. Inductive thematic data saturation was achieved by ensuring that no more new codes or themes were identified during analysis. This shows that majority of the codes were recognized during the first interviews, followed by decreasing codes identified from the rest of the interviews.

### Ethical considerations

The research protocol was reviewed and approved by the Research Ethics Committee of Mbarara University of Science and Technology (Ref: MUREC# 22/09-18). The WHO guidelines on “Ethical and safety recommendations for research on Intimate partner violence against women”^(53)^ were followed. Study methods were executed in agreement with the guidelines and regulations of the Declaration of Helsinki to uphold ethical standards and respect for the respondents to ensure their safety and protected their health and rights ^(54)^. Informed Consent was sought from all study respondents for individual interviews at the time of data collection, enrolled pregnant women of 15 to 17 years of age; need for consent was waived by the research ethics committee and were considered as emancipated minors. The participants were informed that no identity would be used so participants were referred to using pseudonyms names. Interviews were carried out in private rooms by female interviewers to minimize under reporting of IPV experiences. A trained counselor was recruited for the study to emotionally support women that had distress, only two sought for support from the study counselor. No cash incentives were provided, only refreshment in form of a soft drink and a cup cake were given to each respondent to prevent hypoglycemia.

### Rigor and trustworthiness of data

Guba and Lincoln’s criteria including credibility, dependability, confirmability and transferability were followed to achieve rigor and confidence in the study findings ^(55)^.

To maintain credibility of the results, the research team conducted training meetings and pilot interviews. Each of the interviewers conducted two pilot interviews to perfect the overall process using the interview guide, time-management and the overall administration of the interviews. The analyzers had prolonged engagement with the data. Also the multidisciplinary background of the research team in midwifery, nursing, public health and health service research enabled us to interpret the findings. These features guaranteed that data collection and content were consistent.

For dependability the study to dispose of individual analyst predisposition, the three team members (KE, NJB and AJB) were responsible for reading the translated scripts, as well as independently coding and generating meaning out of the data. The individual data was then compiled by the three analysts and evaluated for consistency. Any analyses that were inconsistent or equivocal were discussed amongst the analysts and interpretations clarified until there was consensus among all the three analysts.

For confirmability the analysis was conducted by the research team who had diverse perspectives to the data interpretation. Each analyst used a separate reflexive journal to record different issues that could have affected the analysis process this contributed to appreciation of collective interpretation of the findings. We also used recorded audios and investigators’ field notes as a form of triangulation to validate the data collected. This enabled us to eliminate potential individual investigators’ predisposition and achieve neutral findings reflecting information from the respondents.

To ensure transferability purposive sampling was employed to form a nominated sample of pregnant women who had experienced IPV and we gave an adequate description of the study site and the respondents

## Results

The reported results are derived from In-depth interviews with pregnant women experiencing IPV. Data from interviews are presented in form of cases with direct quotations from the respondents and pseudonyms have been used hereunder.

### Characteristics of respondents

Majority of the participants (56%) were aged 15-25 [range 19 -34] years. Some 64% were in the second trimester, 40% prime gravidae, 40% had small businesses (including tomato and charcoal selling) while 56% had attained secondary level of education, most participants were married (n=92%), the remaining 4% abandoned by partner after awareness of conception, the other 4% runaway after conception due to unbearable violence as shown in table 1 below.

**Table 1.** Characteristics of the participants (n=25)

| Variable | Category | Frequency (%) |
| --- | --- | --- |
| Age(Years) | 15 – 25 | 14 (56) |
|  | 26 – 35 | 11 (44) |
| Trimester | 1<sup>st</sup> | 3(12) |
|  | 2<sup>nd</sup> | 16(64) |
|  | 3<sup>rd</sup> | 6 (24) |
| Gravida | 1 | 10(40) |
|  | 2 | 5(20) |
|  | 3 | 6(24) |
|  | ≥4 | 4(16) |
| Occupation | Salaried job | 6(24) |
|  | Small scale business | 10(40) |
|  | Housewife | 5(20) |
|  | Others | 4(16) |
| **Education level** | No formal education | 1(4) |
|  | Primary level | 4(16) |
|  | Secondary level | 14(56) |
|  | Tertiary level | 6(24) |
| Marital status | Married | 23(92) |
|  | Not married | 2(8%) |

### Themes

Responses from the participants were categorized into four major themes: IPV experiences, support seeking, coping strategies and with intergenerational continuation of IPV. Within each theme, multiple sub-themes were generated to explain the phenomenon better.

### Theme one: Various experiences by pregnant women

Participants’ narratives highlighted various experiences of IPV which were categorized as: Psychological, physical, sexual and economic violence.

### Psychological violence

This was experienced in different forms that include: talking rudely, accused of extramarital affairs, partner receiving phone calls late at night, spending nights outside home, abandoned after conception, refusal to accompany women while attending ANC, chased with a crying child at night and Children witnessing the violence.

Intimate partner talking rudely was commonly voiced by pregnant women in the study.

This made pregnant women sad and frightened by the perpetrator.

> *When we are in the house he can talk to me with a loud voice, it makes me feel bad and I get scared of him”* (**Maureen**).

> *“He talks to me with a loud voice and he doesn’t care whether I get angry or not”* (**Beatrice**)

Most of the pregnant women were accused by their intimate partners of having extramarital affairs and had a tendency to get angry when they found other men talking to these women even in public places. Some were accused of having extra marital affairs with brother in-laws as long as women were out of the intimate partner’s vicinity. This caused psychological trauma since some of these women would be in their gardens toiling for the family welfare.

*He thinks I have other men yet*, *I have a shop most of my customers are men but when he finds me talking to them he becomes angry”* **(Annette).**

> *“He gets mad at me when he finds me talking to other men and wants me to explain what we were talking about, imagine that*!”**(Meron)**

> *“My husband remains in bed, after he accuses me to have sexual relations with his brother yet I’m always in the garden working with his mother”* **(Rehema).**

Intimate partners receiving phone calls late in the night was a common occurrence among pregnant women, claiming that their partners talked for a longtime and were very much suspicious that it were other women they communicated to. This made the victims uncomfortable.

> *“He receives phone calls at night from other women and he talks for a very long time, it annoys me”* (**Annette**)

> *“He receives phone calls late in the night almost every day, I have complained most of the times but he cannot listen to me, it bothers me**”*** **(Godwin)**

Some women voiced that their partners spent nights outside their home without prior notification. This brought mixed feelings of insecurity and fear, wondering if there is another woman and sometimes questioning if he is not in any danger. Some women were told by their intimate partners that since they own the home they can always come back home anytime they wished and women shouldn’t intervene in anyway.

*“Sometimes he does not spend nights at home, he does not tell me where he is, it keeps me imagining a lot of things…..”* **(Imelda)**.

> *Most of the times he comes home towards morning, I cannot tell where he is always coming from, when I ask him he tells me that it’s his house and he can come back anytime he wants**….**bursts into tears* **(Beatrice)**

Some women revealed that the intimate partners abandoned them, blocked their telephone numbers after discovering about the pregnancy. This was often preceded by convincing the women to abort since the partners were not interested in the pregnancy.

> ***“****My partner impregnated me, when I told him that am pregnant he told me to abort I refused, and then he abandoned me and blocked me on his phone right now I can’t tell his where about”* **(Imelda).**

> *At first everything was fine, but when I told him that I’m pregnant his behaviors changed, he told me to abort and I refused, one night he strangled me like he wanted to kill me, I managed to run away and never went back…”* **(Irene)**

Pregnant women expressed concern about their intimate partners failing to accompany them as they seek health care. Some voiced how their intimate partners refuse to escort them for antenatal care attendance claiming that it’s a waste of time.

> *“He refuses to escort me to the health facility that it’s a waste of time for example when going for Antenatal care”* **(Sarah).**

> *“I wish he can accept to accompany me when I’m coming here, but he cannot he always asks me if he is the one carrying the pregnancy”* **(Grace)**

Women being chased out of the house with their crying children were a common form of psychological violence. Intimate partners had a tendency of telling the women to move out of their sight claiming that the children made noise that interrupted their peace and sleep. This made pregnant women feel bad since they expected their partners to first understand why the baby is crying and get involved in child care.

> *“He tells me to carry away my child at night, that he doesn’t want noise ”* **(Rehema,)**.

> *“When any of the children cries at night, he tells me to get off his sight with the crying child as if the child belongs to alone, it hurts!”***(Jackline)**

Some women reported their children witnessing the violence and had nowhere to take the children to prevent them from witnessing the IPV incidences. This brought a lot of discomfort.

> *“What pains me most is that our child witnesses us insulting each other and fighting so often I don’t know where to take the child”* **(Sarah)**

> *“He talks to me any how even the presence of our children”* (**Maureen**)

### Sexual violence

This was experienced in form of Uncomfortable sexual intercourse position and forced sexual intercourse.

Uncomfortable sexual position was one of the mentioned forms of sexual violence. Women claimed that their intimate partners enjoyed positioning themselves on top without caution and not listening to pregnant women’s grievances, yet it’s very painful and uncomfortable due to the gravid uterus.

> *“Laughs….my husband wants to be on top of me during sexual intercourse yet am pregnant and I feel a lot of pain, I have told him that it’s not comfortable but he does not listen he does it every day and it hurts me”*. **(Rose)**

Some respondents reported forced sexual intercourse by their partners, especially after after being beaten by the intimate partner, women became annoyed, lost feelings and interest in coitus yet their partners pretended like nothing had happened and forced them into sexual intercourse. Respondents had no choice but to let their partners fulfill their sexual desires. There was also a common perception that a husband had the right to sexual intercourse at any time regardless of the women’s choice.

> *“Like he beats me, I get annoyed so I cannot have feelings for sexual intercourse, but he tells me to turn and he forces me, I have no choice since he is my husband but when there are no feelings and i feel it on my heart that I don’t want to”* **(Doreen)**.

> *“My husband has bad behaviors, I come from the garden very tired and he forces me to have intercourse with him there and then yet he spends the whole day in bed”* **(Rehema).**

### Physical violence

### This was experienced in form of being slapped, pushed, kicked, stepped on the abdomen and hit

Women expressed concerns of being physically violated on a daily basis while pregnant. This sometimes left women with serious bruises.

> *“… Am now pregnant but he beats me almost every day. He slaps, pushes, kicks and hits me with his fists most of the time”* **(Joan)**.

Some of these fights emanated from refusal of the women to follow the husband’s commands, Rose was not spared by her intimate partner after failing to listen to orders of carrying a bunch of bananas commonly known as “Matooke” from the banana plantation. She knew pregnancy consequences of carrying heavy loads. She wished her husband was considerate such that he can perform the heavy duties by himself. Rose obeyed after a slap in fear of being hit again.

> *“He slapped me on the back during this current pregnancy, he wanted me to carry a bunch of matoke from the banana plantation……”* **(Rose).**

Doreen narrated how she was beaten by Robert her husband and reported that the violence escalated every time she conceived. The worst occurrence was stepping on her abdomen during the fight. Robert always ensured that he left marks on Doreen’s face. The couple already had children and Doreen claimed that she stayed for the children since she had no job and had nowhere to go with them. Doreen claimed Robert had another woman.

> *“…….My husband beats me so badly, especially the time I’m pregnant, he recently stepped on my belly. Every time he beats me he ensures that he leaves a mark on me I can’t go anywhere with these children”*. **(Doreen)**

Marion recounted how she is always slapped on the face, ears and head. To her, most of the times her partner was interested in damaging her face. All this created bad feelings and anger.

> *“My husband slaps me on the face, ears and head. I feel so bad and I’m always very angry**”*****. *(Marion)***

Meron voiced how her intimate partner fought and behaved like a drunkard every day. The fights would be triggered by trivial issues.

> *“My husband is a fighter … he slaps me every day he does things like for a drunkard yet he does not drink any alcohol”*. **(Meron)**

### Economical violence

Economic violence was reported by the respondents and they expressed how their partners stopped them from working to earn money, not given money to seek health care, failure to buy food at home and non-involved in purchase of family assets.

### Lack of financial support

Lack of financial support was one of the forms of economic violence reported. Olivia claimed that her intimate partner had money but refused to pay money for rent, school dues for the children and failed to buy food for the family yet he wanted to eat at home.

This left Olivia stuck with no constructive solution but to tolerate him as the father of the children and man of the house.

> *“You can see that he has money but does not pay for us rent, school fees and food, yet in the morning he wants to eat….. (Cries), but I have nothing to do he is the father of my children… ”* **(Olivia)**

Sharon narrated how Peter her husband wasn’t leaving any money at home for food and other home expenses most of the times Peter claimed that he had no money yet he came back home late and drunk on a daily basis and this kept Sharon wondering where Peter got the money to buy alcohol. She always dedicated her partner to God with hope to change into a responsible man.

*“He always leaves no money and food at home, he comes back towards morning drunk. I keep praying and dedicating him to God with hope that he will change”.* **(Sharon)**

Women expressed their concern about how their intimate partners failed to provide money for transport to seek for medical care and Antenatal care attendance. Sometimes intimate partners ignored these women when they were sick. Women required money for transport and money for food since there are long waiting queues at the facilities. Women believed that this highly contributed to delay in seeking ANC services and sometimes missing appointments.

> *“My husband cannot give me care, like i want to go the health facility, he cannot give me transport and money to buy something to eat on the way so I have to look for it and that is a big problem ……, i buy each and everything by self”.* **(Teddy)**

> *“I tell him am sick but he takes it easy like am not sick he ignores me without giving me money to go to the health facility……., I look for fees for the children and yet I have a partner it bothers me ….”.***(Doreen)**

### Women as sole family breadwinners

Some women claimed to be sole family bread winners and where their main source of income was small scale farming to enable women cater for some basic needs like soap and food. Majority were bothered because they provided everything for the family without intimate partner’s contribution. Women expressed bad feelings about this and admitted that it’s not an easy situation.

*I usually do some small farming to cater for all the basic needs in the home, he provides nothing.”* **(Rehema)**

> *“I usually plant beans when the season is good I sell them and I get some money for basic needs at home, he doesn’t give us anything.”***(Merab)**

### Deprived of gainful employment

Norah narrated how she was stopped from gainful employment by her intimate partner despite the fact that she attained formal education. This was painful and quite disturbing because Norah’s spouse was not providing all her financial needs. She had a perception that what kept the relationship going was the love she had for her partner.

> *“I studied but my husband does not want me to work yet he cannot give me all I want. Since I love him I just bear and I receive whatever he gives me but I keep on having bad thoughts”***(Norah)**.

### Taking away all the money

Olivia described how her partner started a shop for her but the most distressing part was that she was never allowed to directly handle and plan for the money generated from the shop. Her partner routinely takes all the money collected without giving her a single penny, he refills the shop and buys home items by himself. Even when paying a visit to Olivia’s parents he decides alone and buys what to carry. These made Olivia feel like a shop attendant not a wife and a shareholder. It was so disturbing to request for all her personal and home use requirements from her partner. Her wish was to be given the freedom to plan for the money or be paid a monthly salary since she had personal problems that needed money.

> *My husband started a shop for me but every time he comes he picks all the money generated I can never handle money, he re-fills items s for himself am like a shop attendant not a wife and as a woman I also have some things that I would want to buy* **(Olivia)**

There was also a tendency of intimate partners taking away all the money even when the business starting capital belonged to the woman. Rose described how she received money from her mother and started a hair dressing salon but her husband Denis was always interested in taking away all the money made. Even when she is bargaining with the customer Denis stands nearby to know how much is to be paid. This would lead to a physical fight if Rose hesitated to hand in the money. It became so much painful when the Rose wanted to seek for medical care and the partner fails to provide money for costs involved.

> *“I have a salon that I started with my own money, but he takes all the money I make, even when I’m bargaining with the customers he is always around to know how much the cost will be…”* **(Rose)**

Non-involvement in the purchase of family assets was voiced. Respondents reported that intimate partners bought property for example land without informing or consulting women. Incidences of hearing about the new developments from the partner’s friends and sometimes relatives were reported. Some women had no answers why this happened, though others attributed it to disrespect since most of these women had no sources of income to contribute to the family’s properties bought. And other thought that their partners could be planning to marry other women.

> *“He buys property without telling me, if I get lucky I hear it from friends it really hurts” (***Emily**)

> *“He buys family property like land without informing me, I think he does not involve me because I have no source of income to contribute money, but I cannot be sure he could be buying it to marry another woman….”***(Joyce)**

### Theme two: Support seeking

Participants were probed whether they sought help after IPV experience and a number of them sought support through informal and formal means, as mentioned in the quotations. Informal support was from respondents’ biological relatives like mothers, siblings, and fathers. A few sought support from their mother in laws and friends. Others never sought support due to lack of whom to confide in and fear of being laughed at.

> *“I usually tell my mother and she counsels me”* **(Miriam).**

> *“I usually tell my father all my experiences sometimes he sends money to support me….When it’s too tough he encourages me to come back home and rest”* **(Bridget)**

It was voiced that spouse’s friends had a tendency to withdraw from the matter after disclosure. Also it appeared that these friends had no experience in handling intimate partner violence issues since they never called back to inquire about the progress in regard to the disclosed violence, some participants felt these friends withdrew to maintain their relationship with the perpetrator.

> *“I told his friend about the violence, he was shocked about everything and promised to talk to him. I expected his friend to call back first and give feedback which he never did, so i called and confirmed that they met and talked, you see it didn’t help much I wish I had kept quiet…..”* **(Godwin**).

Some respondents never sought support due to lack of trust and others reported to the human rights organizations and others to police depending on the severity of IPV and some observed change thereafter.

> *“If he beats me every day I report to the police or human rights organizations such that he is threatened and sometimes counseled”* **(Lydia)**

> *“Look at the scar on my face, I sustained it when he hit me hard and I reported him to police and he later changed his behaviour”* **(Miriam)**

### Barriers to support seeking

Some women never sought support due to trust issues, threats from the perpetrator and fear of the perpetrator’s knowledge about any support sought.

Some respondents believed that those they sought help from would spread the rumors to non-significant others that would instead laugh at them. Some found it difficult to identify people they trusted to provide support with some reporting that they had no biological mothers which made it difficult to identify someone to confide in thus waiting patiently for the husband to change.

> *“I look for whom to tell …laughs. Anyone I tell will laugh at me and will go telling others that I was beaten so I decide to keep quiet”* **(Marion).**

> *“I have no mother, I therefore keep my problems and I try to talk to my husband such that he can change”* **(Teddy).**

Some received constant threats from the violent partner who warned not to tell anyone about the ongoing violence claiming that they would abandon the women with the children and never to back. This controlled the women since majority were housewives with no gainful income to financially support the children.

> *“He threatens me that when he beats me or mistreats me and I go to police or tell any other legal authorities he will abandon me and never to come back, I cannot afford the finances to care for my children so when I hear that I cool and leave everything”* **(Emily).**

Others feared that their husbands would become more violent after knowing that a third party had been involved. Some had experienced the worst violence after involving someone else in these family issues. This restricted respondents to only seek support from biological mothers they fully trusted, with surety that the perpetrator won’t know about the support sought.

> *“One time I told my brother in law about our problems in the home, when he talked to my husband about it, the violence increased! That night he beat me up terribly, he strangled me like he wanted to kill me. So I can now only tell my mother since I know the information will not get back to him ”* **(Sarah)**

### Midwives’ involvement

Participants were further probed to know if they sought support from midwives. Surprisingly none had ever done so due to different perceptions. Some women wondered how to start involving health care providers in marital issues. Participants generally thought that IPV is not a health care provider’s role. To them health care workers were supposed to measure their blood pressure, perform abdominal examination and provide treatment and they voiced that healthcare providers never inquire about IPV:

> *“I wonder how I begin telling her. Now how do you begin? ……That’s not her work! “***(Jackline)**

> *“Their work is to educate us about diet, what to bring to the hospital at the time of delivery* **(Annette)**.

> *Why should she even ask me how my husband is treating me! Is she my aunt? I have come to check for pregnancy. “*(**Beatrice).**

> *“Sometimes I feel like telling a midwife but I wonder where to start from. She has not asked how things are at home I have come for antenatal care and I start saying problems at home no….***(Norah).**

> *If she asks me then I can get where to start from but they speak to us rudely.”* **(Merab)**

> *“You are the first one I have seen asking me”* **(Godwin).**

> *“Even you, I wouldn’t have told you if you had not asked me.*

> *They don’t ask…they also have their own problems and they start asking about my own problems instead they will put their stress on me”* **(Imelda**).

### Theme three: IPV coping strategies among pregnant women

Participants experiencing IPV during the index pregnancy mentioned coping strategies including: confiding in their relatives especially mothers of the victim, a few to mother in-laws, keeping quiet, self-consolation, partner tolerance, reporting to authorities, disrupting negative thoughts, praying to God, state of resignation, consideration of man as a dominant figure, understanding spouse’s likes and dislikes, saying yes to everything and belief that all men are the same.

### Confiding in relatives

The most common confidants were the biological mothers of the victims who often encouraged their daughters to stay in the relationship despite the IPV incidents. Most of them were advised by their mothers to always keep quiet as a way of creating a peaceful situation when their partners started quarreling. To these women talking to a confidant like a mother relived stress.

> *“……my mother always tells me to keep quiet and ignore him.”* **(Godwin)**

> *“When I had problems with my husband, I talked to my sister she counseled me and told me to be strong and patient and indeed I became strong and managed”* **(Joan).**

A few like Rehema confided in their mother in laws. Rehema was close to her mother in law and they would do gardening together this made it easy for her to share her experiences, Rehema’s mother in law encouraged her to be patient giving her hope of success in future.

> *“I first told my mother in law about the violence, she told me to be patient”* **(Rehema).**

### Keeping Quiet

Doreen revealed that she keeps quiet and pretends to be a fool in response to the violence caused by her partner. Doreen’s perception was that keeping quiet prevents escalation to physical violence.

> *“Most of the times he quarrels and I pretend to be a fool so I always keep quiet to avoid slaps and punches…”* **(Doreen).**

Though there were mixed feelings where by some women benefited from silence while others did not. Marion narrated that sometimes she tried to keep quiet but her husband instead became more furious complaining that Marion was stubborn due constant silence.

> *“Most of the times I keep quiet though that makes him more furious that I’m stubborn and bigheaded and damn, hmmm it’s not easy**.”***(Marion)

Others felt that their opinions were not valued by their intimate partners hence keeping quiet was the best solution.

> *“Even if I argue he will not value my opinion so I just keep quiet**”*** **(Joyce). Praying to God**

Respondents also coped by getting closer to God through meditation and prayer when IPV occurred. Others approached their Pastors to pray for them.

> *I always pray to God and my pastor prays for me* **(Jackline).**

> *I usually dedicate him to God, When you keep on praying God changes him…….* **(Grace).**

### Disrupting negative thoughts

Some coped through disrupting negative thoughts and replacing them by thinking about other good things like friends to have a piece of mind.

*“I try to think about other good things like my friends such that I take away all the bad thoughts.”* **(Joyce)**

### Reporting to authorities

Reporting to authorities was one of the adapted coping ways. Women believed that their husbands respected the local council, non-governmental organizations and the police.

> *“My husband hit me hard……that I didn’t want to give him a child, thank God I had conceived though we hadn’t known. I reported him to Police”* **(Bridget).**

### Self-consolation

Due to cultural concerns, public opinion towards marriage and public respect accorded to married women there was a tendency of coping through self-consolation. For Sarah’s perception was that in their culture a woman without a spouse is undermined and disrespected by the community. To maintain this perceived respect and social reputation, Sarah consoled herself and stayed in the violet relationship.

> *I console myself with the public picture for the respect of being married….. Many people around will not respect me when I have no husband”* **(Sarah).**

Despite the intimate partner’s violet behaviors, others consoled themselves when the partner was constructive in terms of home building, paying school fees for the children and providing food for the family. This was a fundamental comfort to remain in the violet relationship. Women under this category felt better than other women in violet relationships whose intimate partners fail to sustain the family financially.

> *“His behaviors are bad but he is constructive, he pays fees for the children and he ensures we have food so I console myself that I’m better than many out there”* **(Meron)**.

### Partner tolerance

Some tolerated their partners despite the IPV experience because of lack of alternatives including financial support and the fear of abandoning her and the children. There was also an element of partner tolerance as long as the intimate partner was still interested in having sexual intercourse. Sexual intercourse influenced the belief that there was still love between them and also dispelled worries of the partner having extramarital relationships.

> *“I have nothing else to do, once he does not want to talk to me but gives me money to buy food and he is willing to have sexual intercourse with me, then I can bear him the way he is…..’’.* **(Irene).**

### Man as a dominant force

A tendency of women considering their intimate partners (Men) as superior was cited. Respondents mentioned that a woman cannot compete with a man since he can easily beat her. To them the solution was keeping quiet when violence sets in.

> *“You cannot compete with a man, if I become stubborn, he can slap me……”* **(Norah).**

### State of resignation

A state of resignation was noted. Some respondents had histories of prior relationships that were violet. This forced some to withstand the violence in fear of being blamed by the society as complicated women and these partly blamed themselves and had to remain in the relationship.

> ***“****I decided to bear the situation because this is my second relationship, my first partner used to mistreat me and I left him….. If I decide to quit now people will judge me as the complicated one”* **(Merab)**

### Understanding spouse’s likes and dislikes

Understanding what the spouse liked or hated was voiced as one of the coping strategy.

> *“I have managed by trying hard to know what he likes and what he hates, hmmm….,”* **(Olivia)**

Saying yes to everything:

> “Whatever he says whether wrong or right I say yes yes…to avoid being beaten” **(Norah)**

### Belief that all men are the same

Some participants believed that all men are them and believed that everyone has a specific burden to tolerate in a relationship.

> *I accepted him the way he is, because it’s not different in other families, everyone has a cross to carry”* **(Olivia)**

### Theme four: Intergenerational continuity of IPV

Intergenerational continuity of IPV was noted, where some of the participants voiced that their Mothers in laws interrelated to how they also went through the same violence with the respondents’ father in laws. This made most of the respondents calm down after realizing that their mothers in laws have gone through such IPV experience.

> *“My mother in law told me to be patient that’s how his father used to treat her! You will succeed one day…”* **(Rehema).**

## Discussion

This study explored the pregnant women’s IPV experiences, support seeking and coping strategies in Southwestern Uganda. We report women’s voices of IPV experience in form of physical, psychological, sexual and economic violence. Women preferred sharing IPV experiences with their biological mothers to midwives or any other person. The main support given by their support systems was encouraging the victims to maintain their marriage and keeping quiet when the partner starts quarreling. Pregnant women did not know the role of midwives in mitigating IPV and the midwives’ did not investigate about the IPV experience during Antenatal care contacts. Women coped by confiding in their relatives, keeping silent, self-consolation, tolerance of the perpetrator, distracting bad thoughts through thinking about good things like friends, self-blame and praying to God and intergenerational continuity of violence was narrated.

### IPV experienced by pregnant women

Current findings reveal that women experienced various types of IPV including psychological, physical, sexual and economic violence. This is comparable to previous studies that reported psychological violence physical, sexual ^(5, 13, 19)^ and economic violence ^(6, 14)^. A number of women expressed their concern on economic violence where they claimed that spouses had money but ignored their financial needs that included paying money for rent, school fees for the children, buying food for the family. This situation can intensify the violence since in African settings men are traditionally considered as family breadwinners. This is supported by previous studies which asserted that women are never breadwinners but have supportive roles in families ^(10, 56)^. In the African tradition it is known that a man should be economically able and willing to provide almost everything to the family including providing for the wife ^(11)^. Studies have affirmed that when women are left to be the main source of family survival, instabilities and divorce may set in^(57, 58)^.

Non-involvement in purchase of family assets and restriction in obtaining gainful employment was voiced by the respondents and others who were allowed to work all the money was taken away from them. This is in agreement with aforementioned studies ^(51, 59–61)^, that economic violence was widespread where violent partners impede women’s employment, access to financial resources and isolation from domestic financial information. This creates economic dependency on the perpetrator, forcing women to beg for everything and become more controlled. Studies have consistently demonstrated that economic dependence is the primary obstacle victims face in attempting to leave violent relationships ^(7, 8, 62)^. Women need to be empowered with personal development skills, education and knowledge on problem solving skills to reframe their situation and regain power from the perpetrator.

Women narrated how their partners insisted on uncomfortable sexual positions specifically man on top commonly known as the missionary style. This is consistent with previous studies ^(63–67)^ where the missionary style remained common position throughout pregnancy. During pregnancy increase in abdominal volume and girth makes this common position more difficult and uncomfortable ^(67)^. Sexual intercourse shouldn’t be abridged to the man’s interest: pregnant woman’s satisfaction, comfort and enjoyment should also be considered. Due to societal norms, women may not freely talk about these sexual issues ^(64, 68)^ and continue with such experience while trying to fulfill marital obligations. Consequently midwives in the prenatal clinics should have the initiative to facilitate, educate and counsel pregnant women and their partners on appropriate sexual positions, particularly after 20 weeks of gestation to keep pressure and weight off the abdomen and positions that keep the woman off the back to avoid potential blood flow compression that can lead to discomfort, fainting and other issues. The recommended sex positions during pregnancy include woman on hands and knees, woman on top, seated pregnancy sex, side by side, use of pregnancy pillows ^(69, 70)^ for support and more comfort.

Intergenerational violence was voiced by the respondents, this is in line with earlier studies ^(71–74)^ that reported experience of violence during childhood contributes to victimization and perpetration during adulthood. This can be attributed to lack of maternal-infant bonding during the period of IPV experience which affects the child’s development and behavior ^(75)^, also the tendency of children demonstrating their parents’ behavior and act the same in their relationships during adulthood. It is therefore important to identify victims early which can enhance timely intervention and minimize on the future consequences.

### Support seeking and midwives involvement

Women preferred seeking support informally from their biological relatives compared to midwives or any other person. The results are similar to previous studies ^(76, 77)^. Women sought support specifically from their birth relatives probably due to the primary bonding, security and the trust they have in their biological relatives. It is reported that women have a strong link with the family of origin ^(76)^. Having somebody to confide in should be credited since it is recounted that lack of victim family support acts as a barrier to terminating IPV ^(78)^. Another reason why women prefer to confide in their natal relatives is because they fear that their intimate partner’s family would support their son and prevent the issue from being resolved. Fear of the perpetrator discovering that a third party was involved is another possibility, since victims normally reside far from their biological relatives, it makes it difficult for the abusive partner to learn about the support being sought. It is reported that fear of the perpetrator negatively affects support seeking ^(79)^. The current findings are contrary to the study by Amel Barez et al ^(80)^ where the bulk of the women never asked for help and kept their experiences with perinatal IPV hidden..

Interviews revealed that the participants in the current study were unaware of the role that midwives play in IPV. The expert opinion in an earlier studies indicate that women are less likely to reveal violence to institutions such as health facilities because they appear to be more formal, and they do not know what formal measures would be taken when health care providers are involved to stop further violence ^(36, 42)^. Nurses and Midwives form the biggest healthcare workforce in Sub-Saharan Africa ^(81)^ and most of the prenatal clinics in these settings are operated by midwives so their role and responsibility in IPV control and management is crucial. As part of this, IPV screenings are conducted, resources are provided to victims, women who sustain physical injuries are treated, the victim’s and significant others’ safety is assessed, the findings are properly documented, emotional support is provided, and appropriate referrals are made ^(75)^. Many women may focus more on the immediate solution to stopping the violence and neglect to consider how IPV impacts them in the long run. Health care providers responsible for maternal and child health, specifically midwives, are encouraged to educate pregnant women about their role in preventing IPV, to elaborate on the consequences of IPV during pregnancy, as well as to actively screen pregnant women for IPV and offer support. By doing so, women will realize and appreciate that care providers play an important role in this regard, and this will help them break the cycle of IPV.

### Coping strategies

Among the coping strategies used by pregnant women in this study were confiding in someone commonly their biological mothers, partner tolerance, self-consolation, self-blame, keeping quiet, thinking of other good things like friends, praying to God, and others reported to authorities. This is comparable with a study done in Bangladesh where in particular, women used different strategic responses, including remaining silent, trying to divert their intimate partner’s attention, protesting against the violence, and seeking help from others ^(82)^. Some of these coping mechanisms for example keeping quiet may shortly calm the situation since it prevents escalation to physical violence. On the other hand silence puts the victim at risk of continuous exposure to IPV due to the fact that it does not encourage dialogue between the partners to solve the conflict. It has been reported that persistent violence in intimate relationships leads to chronic stress, which survivors describe as painful and overpowering, posing a threat to their physical and social self ^(83)^. IPV results in significant morbidity and disability among women with the biggest impact on their reproductive and mental health ^(84)^. Traumatic stress is thought to be the main mechanism that explains why exposure to intimate partner violence may cause subsequent depression and suicide attempts to the mother as previously reported ^(85)^. This also affects the fetus. The fetal effects include: preterm delivery, low birth weight, fetal death ^(34, 86)^ and if the baby survives with continuous IPV exposure, is highly likely to be at risk of adult victimization and perpetration ^(74, 75)^.

Findings show that women resorted to praying to God and involving religious leaders to pray for them. This is echoed in previous studies ^(43, 44)^. Not different from a previous study ^(87)^, in the current study women hoped that their intimate partners would change if they continued praying for them. It has been documented that religious coping can benefit the victims through reshaping their perceptions and institute new behaviors, and can support recovery by providing hope and preventing feelings of powerlessness ^(88)^. Though too much devotion in prayer can at times be dangerous since some women may go to the extreme of spending much of the time in churches praying including overnights, others stop working and spend all the time praying which can worsen the relationship with their partners due to long periods of absence at home. Women voiced reporting IPV experience to authorities like police when IPV became severe remarkably associated with injuries. This is in agreement with reports from previous studies which showed that victims sought for help when the IPV experience was severe and could not endure anymore ^(37, 76)^. This indicates that some women can tolerate IPV when it’s not severe and life threatening which is not healthy since any level of IPV has significant consequences to the women, fetus, children and other family members.

## Conclusion

Pregnant women continue to live in violent relationships with various coping strategies and women are not aware of the midwives’ role in IPV. These findings point to the need for the health care system to reflect on the care provided to these women in our health centers, community clinics and hospitals in response to women who are not aware that family IPV can be discussed with health professionals. Officers in charge of maternal and child health need to develop and incorporate a user friendly IPV screening tool onto the ANC card, ensure midwives and obstetricians professional training on the process of IPV identification and management, mentoring and supervision to enhance routine IPV screening, also recruit counselors and peer support specifically for pregnant women in the prenatal clinics to provide individualized psychological support. Professional health education talks on sexual positions during pregnancy need to be emphasized during ANC classes and community outreaches to prevent sexual violence in regard to uncomfortable positions. Policy makers in maternal and child health department should develop a protocol for care of women identified to have suffered from IPV, ensure follow up mechanisms for continuum of care.

### Study strength and limitations

One of our strength was achieving data saturation by interviewing a sample of 25 women experiencing IPV and attending ANC. The second strength was using qualitative methods of inquiry which are the best in understanding pregnant women’s experiences and coping strategies. Also existing literature in our setting majorly considers psychological, physical and sexual violence, the current study reveals economic violence too. This study had some limitations. First the findings may not be transferable to rural health facilities since our study was based in the urban setting. Secondly this study relies on self-report, so recall bias and inaccuracy may be present. Nonetheless it contributes literature on IPV which is still a very fundamental topic. Future research should focus on triangulating views from rural health facilities and urban health facilities to appreciate the diverse views that can diminish the possible bias created by this concept.

## Data Availability

The data used to support the findings of this study are available from the corresponding author upon reasonable request.

## Acknowledgement

The authors extend their gratitude to the First Mile Community Health program for the support during the time of the study. Great thanks to the Department of Community Health, Mbarara University of Science and Technology for the support rendered during the study. The authors would also like to acknowledge the study participants for their willingness and cooperation during the data collection process.

## Author’s contribution

KE, NJB and BV designed the study. KE participated in data collection. KE, NJB and AJB participated in data analysis. KE, NJB, AE, KA, NG, AJB and BV participated in original drafting, review and editing of the manuscript. All authors read and approved the final manuscript.

## Consent for publication

Participants gave their consent for the data to be used for research purposes. They were also assured that any information about them will be anonymized.

